# A cross-tissue splicing signature as a quantitative biomarker for ReNU syndrome

**DOI:** 10.64898/2026.09.26.26364077

**Authors:** Ruebena Dawes, Alexander Blakes, Theodora Markati, Michael Griffiths, Benjamin Cogné, Tahsin Stefan Barakat, Jeffrey C Barrett, Carlo Rinaldi, Stephan J Sanders, Nicola Whiffin

## Abstract

ReNU syndrome is a severe neurodevelopmental disorder caused by *de novo* variants in the spliceosomal small nuclear RNA (snRNA) gene *RNU4-2.* Pathogenic variants cluster in two distinct structural regions of the U4 snRNA produced by *RNU4-2*, the T-loop and Stem III. Aberrant 5’ splice site selection is a molecular hallmark of ReNU syndrome that correlates with phenotypic severity and could be leveraged as a quantitative biomarker to facilitate preclinical and clinical therapeutic development. We analysed RNA-sequencing data from two independent cohorts totalling 30 individuals with ReNU syndrome and 54 controls. We identified 483 alternative splicing events shared across both cohorts, with the highest concordance in 105 alternative 5’ splice-site (A5SS) events. Using these 105 A5SS events, we derived a minimal reproducible signature from only six sites. The splicing signature perfectly distinguished 19 n.64_65insT individuals from 54 controls, and produced no false positives when applied to a further 5,984 whole blood controls. In addition, the signature was specific to variants within the U4-2 T-loop, which are associated with greater clinical severity. Further, we find evidence of alternative splicing events that are specific to variants in Stem III. In a patient-derived induced pluripotent stem cell (iPSC) model of the recurrent *RNU4-2* n.64_64insT variant, the splicing signature was fully recapitulated across all stages of differentiation into cortical neuron-like cells. Together, these findings define a robust RNA biomarker for T-loop ReNU syndrome for use in therapeutic development, and illustrate a general framework for portable biomarker discovery in spliceopathies.

## Introduction

ReNU syndrome is an autosomal dominant neurodevelopmental disorder (NDD) caused by *de novo* variants in *RNU4-2* that may affect ∼100,000 individuals globally^1,2^. The *RNU4-2* gene is transcribed to U4-2, the predominant isoform of the U4 small nuclear RNA (snRNA) that is a key catalytic component of the major spliceosome. Variants that cause ReNU syndrome are confined to two structural regions of U4-2, the T-loop and Stem III regions, within an 18 nucleotide (nt) contiguous region of the 145 nt long RNA^1,2^. A single variant within the T-loop, n.64_65insT (NR_003137.3), accounts for 70-75% of all ReNU syndrome diagnoses^1,3^.

Within complex B of the major spliceosome, the U4 snRNA tethers the U6 snRNA to allow highly specific 5’ splice site selection by the U6 ACAGAGA motif^4^. Consistent with this known function, pathogenic variants in ReNU syndrome result in widespread disruption to 5’ splice site usage^1^. The extent of this splicing disruption has been shown to correlate with phenotypic severity in both patients^3^ and cellular models^5^. The majority of aberrant 5’ splicing events detected in individuals with ReNU syndrome involve annotated rather than novel alternative 5’ splice sites^6^, suggesting that variant U4-2 may result in reduced specificity for canonical splice sites.

Given the severity and prevalence of ReNU syndrome, and the preponderance of a single causative variant, *RNU4-2* is an attractive therapeutic target. The splicing changes associated with ReNU syndrome could be used as a quantitative biomarker to accelerate pre-clinical and clinical therapeutic development and monitor treatment efficacy. This approach has precedent in other monogenic diseases^7^. However, RNA sequencing (RNA-Seq) experiments are highly sensitive to tissue- and batch-specific effects^8,9^, and need to be validated across model systems and sequencing methods. Therefore, a major challenge is to ensure biomarker robustness and reproducibility across different contexts.

Here, we develop a splicing signature and RNA-based biomarker for ReNU syndrome. Using RNA-Seq from two independent cohorts, we identify concordant disease-associated alternative 5′ splice-site perturbations among individuals with the n.65_65insT variant. We derive a minimal six-event splicing signature that accurately distinguishes ReNU individuals from controls across tissues and experimental systems. Together, our findings establish a robust molecular readout of spliceosomal dysfunction for therapeutic monitoring and illustrate a general strategy for RNA-based biomarker discovery in spliceopathies more widely.

## Materials and methods

### RNA sequencing cohorts

RNA sequencing data were analysed from two cohorts comprising individuals carrying pathogenic *RNU4-2* variants and unaffected controls (Figure 1A).

**Figure 1:**
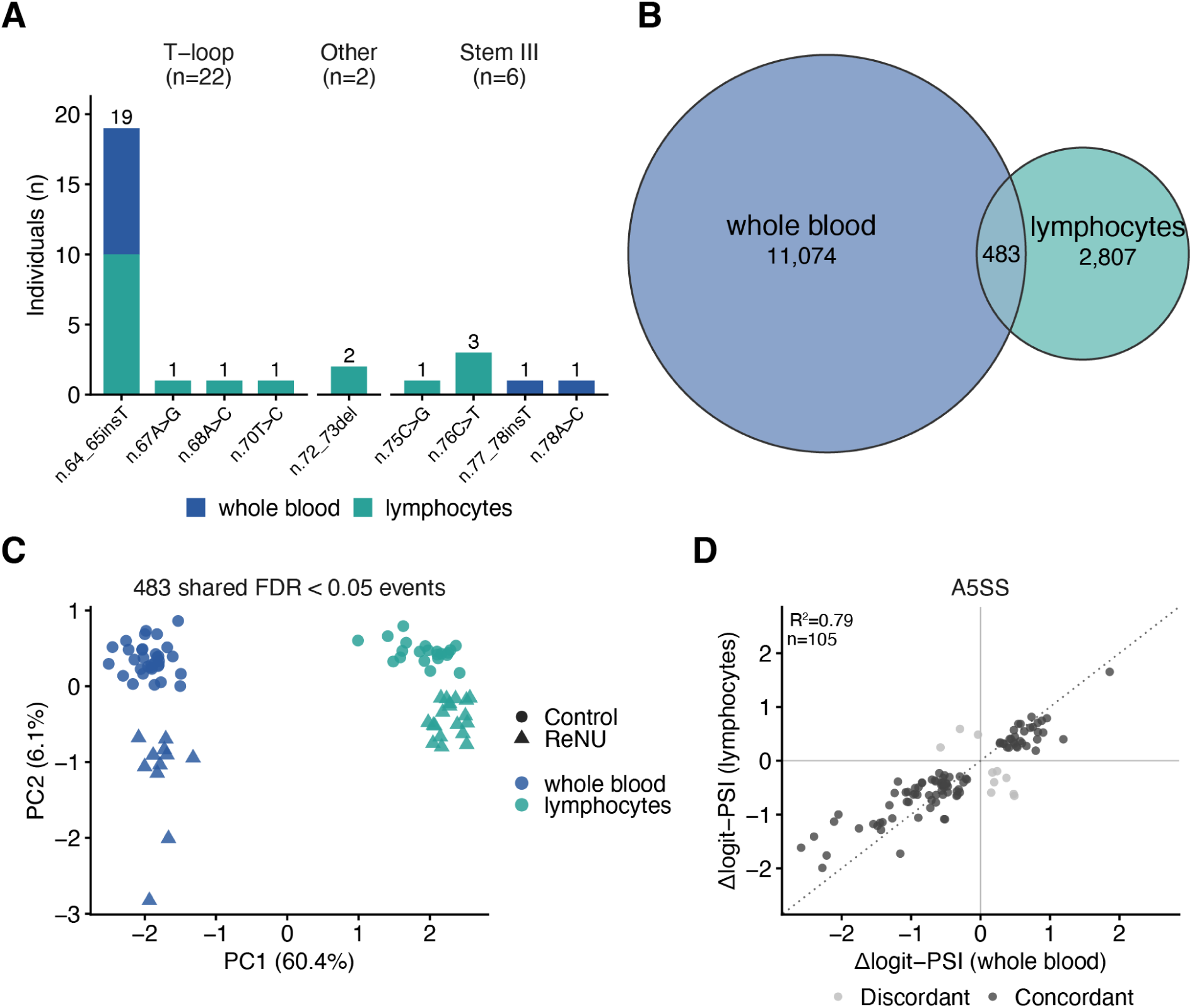
Cross-cohort analysis of ReNU syndrome associated splicing changes. **A)** Distribution of *RNU4-2* genotypes in the two ReNU syndrome RNA-seq cohorts. **B)** Venn diagram showing significant alternative splicing events detected by rMATS^20^ in whole blood and lymphocyte cohorts (false discovery rate (FDR) < 0.05), among the set of events which could be quantified in both cohorts. **C)** Principal component analysis (PCA) of percent spliced in (PSI) values for the 483 alternative splicing events that were significantly differentially spliced (FDR < 0.05) in both the whole blood and lymphocyte cohorts. **D)** Scatterplot of the mean Δlogit-PSI values across ReNU individuals versus controls in whole blood versus Lymphocyte datasets for 105 alternative 5’SS (A5SS) events significant in both cohorts. Points are coloured by whether the effect direction is concordant (dark grey) or discordant (light grey) between cohorts.

#### Development cohort 1 (whole blood)

Whole blood ribodepletion RNA-seq samples generated within the Genomics England (GEL) 100kGP Transcriptomics pilot and extension (https://re-docs.genomicsengland.co.uk/rna_seq/). Read alignment and transcript quantification were performed using the DRAGEN RNA Pipeline v.3.8.4 and v.4.2.7, with annotations from gencode v.32. Among the 7,829 probands with RNA-Seq within Genomics England, 12 ReNU syndrome patients were previously identified^10^. One ReNU sample which was an extreme outlier in total read counts as noted in the Genomics England documentation (https://re-docs.genomicsengland.co.uk/rna_seq_pilot/) was excluded from splicing signature development.

For each of the 11 remaining ReNU syndrome samples, we selected three control RNA-seq samples matched on age at consent, sex, ancestry and mil mapped reads. Controls were selected from a set of 7,829 RNA-seq samples in Genomics England with mil mapped reads > 60 and excluding individuals with normalized disease group = ‘neurology and neurodevelopmental disorders’.

#### Development cohort 2 (lymphocyte)

The lymphocyte cohort was derived from a published study where ReNU patients were identified among a cohort of 15,073 patients with NDD and their parents in France^3^. Briefly, the RNA-Seq samples in this cohort were Lymphocyte RNA-seq samples obtained from peripheral blood mononuclear cells, from 19 ReNU patients and 21 control samples matched on library preparation kit, sequencing flow cell and culture time.

Across both development cohorts, alternative splicing was quantified using rMATS-turbo (v4.3.0) with paired-end, stranded settings (--libType fr-firststrand), –anchorLength 1, and novel splice-site detection enabled (--novelSS). The whole blood cohort used the GENCODE v49 annotation set while the lymphocyte cohort used Ensembl Release 112. For both cohorts the rMATs Junction Count output was used.

#### Larger control cohort (whole blood)

RNA-Seq samples from the 7,829 probands within GEL were filtered using RNA-seq quality-control thresholds previously applied to the 100kGP Transcriptomics pilot^11^, These filters were: total reads ≥120 million, estimated uniquely mapped reads ≥30 million, mapping rate ≥95%, and rRNA rate ≤12% using the rnaseq_qc_metrics Labkey table. Uniquely mapped reads were estimated as:

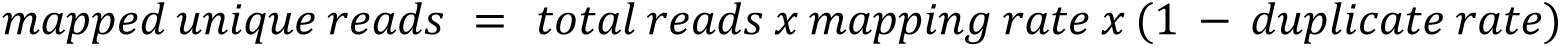

These QC filters, along with excluding ReNU samples and matched controls used in the development cohort, as well as the high-depth ReNU case and individuals with known biallelic *RNU4-2* variants, resulted in a set of 5,984 samples.

Alternative splicing quantification was performed using rMATS-turbo (v4.3.0) partitioned into batches of 100 samples in --statoff mode against the GENCODE v49 annotation set with paired-end, stranded settings (--libType fr-firststrand) and novel splice-site detection enabled (--novelSS).

### Splicing Signature Model Construction

For each alternative splicing event found to be significantly altered in ReNU individuals (FDR < 0.05) in both the whole blood and lymphocyte cohorts, we transformed rMATS PSI values using a pseudocount-stabilized logit transformation:

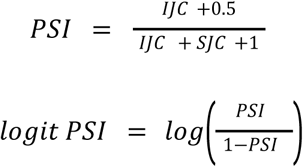

The logit transform gives small absolute PSI changes near 0 or 1 weight comparable to changes in the middle of the range, which has been used to better represent splice-site changes as competition between alternative splice-sites^12,13^. We then calculated Δlogit-PSI values by calculating mean control logit PSI values (µ*_ctrl_*) for each event separately per cohort, and calculating the Δ for each ReNU sample versus their cohort µ*_ctrl_* like so:

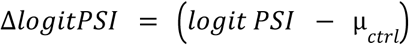

For model training, candidate splice events were restricted to alternative 5′ splice-site (A5SS) events quantified in both cohorts satisfying all of the following criteria: false discovery rate (FDR) < 0.05 in both whole blood and lymphocyte cohorts, concordant direction of Δlogit-PSI between cohorts, and minimum junction depth ≥10 reads in every case and control sample, where depth was defined as rMATS-derived Inclusion Junction Counts (IJC) + Skipping junction counts (SJC) for the event.

Whole blood and lymphocyte individuals with n.64_65insT were each partitioned randomly into training (70%) and holdout (30%) sets to ensure both cohorts were used for training. For each splicing event, sample-level deviations were multiplied by either 1 or -1 such that ReNU-associated deviations were positive. To reduce redundancy arising from multiple splice events within the same gene, a maximum of one event per gene was retained prior to model fitting. Events were ranked using the geometric mean of Mann–Whitney-derived AUC estimates (*G_AUC_*) in whole blood and Lymphocyte cohorts for differentiating between cases and controls:

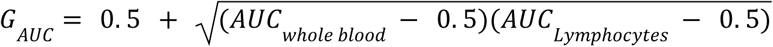

The highest-ranked event per gene was retained for downstream modelling.

A logistic-regression classifier was then trained with L1-penalized regression. Model fitting was performed on the feature matrix of *x_ij_* for 77 events (*j*) in 68 samples (*i)* (14 training cases and all 54 controls from both cohorts). Non-negative coefficient constraints were imposed such that all retained features contributed positively to disease classification. The regularization parameter λ was selected according to the λ_1*se*_ ,defined as the largest λ value whose mean cross-validated deviance (disagreement between predicted case probabilities and true sample status on held-out samples) was within one standard error of the minimum.

The final classifier selected six A5SS events. Predicted probabilities of case status were generated according to:

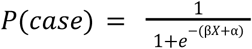

where *X* denotes the feature matrix restricted to the 6 selected events, β the fitted coefficient vector and α the intercept term.

We tested for overfitting by repeatedly selecting sets of 19 pseudo-cases and 54 controls randomly from among the 5,984 whole blood controls. For each permutation, we ran rMATS –task stat to find significantly altered A5SS events in the pseudo-cases (FDR < 0.05). As in the development of the genuine splicing signature we then filtered the events to those with minimum junction depth ≥10 and the highest-ranked event per gene. We randomly split the pseudo-cases into 14 training and 5 holdout cases, attempted to fit a LASSO model with λ_1*se*_ , and reported the AUC in both training and holdout samples across 1,020 permutations.

We derived a stem-III model by replicating the process, with two differences: (1) cases were the six individuals with Stem-III variants rather than insT individuals, and (2) because of the small sample size, no train/holdout split was used.

### iPSC lines & culturing

We used three independent clones from a single iPSC line derived from skin-derived fibroblasts previously obtained during routine diagnostics from an individual with a *de novo RNU4-2* n.64_65insT variant. iPS reprogramming occurred following established standard procedures at the Erasmus MC iPS facility^14^ using the Cytotune 2.0 Sendai reprogramming kit. Details on the individual were previously published^15^. As control, neurons derived from three independent passages of the well-established reference iPSC line KOLF2.1J^16^, from now on referred to as KOLF (which is also derived from fibroblasts) were used.

Human iPSC lines were maintained in mTeSR Plus (Stemcell Technologies) medium on hESC-qualified Matrigel (Corning)-coated 10-cm dishes. Cells were passaged at ∼80% confluency using 0.5 mM EDTA (Thermo Fisher) in PBS for detachment, followed by centrifugation at 200 x g for 5 min at 25 C to pellet the cells. For the first 24 h after passaging cells were maintained in mTeSR Plus medium (10 ml for 10-cm dish) and 10 μM (1:1000) Rho-associated protein kinase (ROCK) inhibitor (RI; Y-27632, Selleck Chemicals) reconstituted as 10 mM in PBS. Cultures were maintained in a humidified incubator set to 37 C with 5% CO2.

### Generation of iPSC lines containing *human neurogenin-2* (*hNGN2*) under a tetracycline-inducible promoter

Generation of iPSC cell lines expressing human *neurogenin-2* (*NGN2*) under a tetracycline-inducible promoter was done using a PiggyBac system for delivery, as described in Pantazis *et al*. (2022)^17^. The plasmids PB-TO-hNGN2 (Addgene plasmid #172115) and EFa1-Transposase (Addgene plasmid #172116) were used for this purpose. PB-TO-hNGN2 was a gift from iPSC Neurodegenerative Disease Initiative (iNDI) & Michael Ward. To generate stable iPSC lines, cells were transfected with 1:2 ratio (transposase:vector) using Lipofectamine Stem (Invitrogen). 48 h post-transfection, puromycin selection was started; cells were selected continuously for 14 days starting at 1 μg/mL and gradually increasing up to 8 μg/mL of puromycin.

### iPSC differentiation to cortical neurons and characterization

Neuronal differentiation of human iPSC lines with a stably integrated human *NGN2* followed a well-established protocol with minor optimizations as described in Fernandopulle *et al*. (2018)^18^. Human iPSCs were single-cell dissociated using Accutase (Thermo Fisher) and plated on a Matrigel-coated 15-cm dish [(Day 0 (D0)] with neuronal induction medium (IM). IM was prepared and changed daily as follows: Gibco KnockOut DMEM/F-12 (Thermo Fisher), 1X Gibco N-2 Supplement (Thermo Fisher), 1X Gibco Non-Essential Amino Acids (NEAA; Thermo Fisher), 1X Gibco GlutaMAX (Thermo Fisher), and 2 μg/mL doxycycline (Sigma-Aldrich) reconstituted in PBS. On D3, cells were replated onto poly-L-ornithine (PLO; Sigma-Aldrich) and laminin (Corning)-coated 6-well plates (for biochemical applications), or µ-Slide 8 Well high Glass Bottom (ibidi GmbH; for immunocytochemistry) with cortical neuron culture medium (CM). CM was prepared as follows: BrainPhys (Stem Cell Technologies) 1X B27 Supplement (Thermo Fisher), 10 ng/mL BDNF (PeproTech), 10 ng/mL NT3 (PeproTech), 1 μg/mL Laminin (Corning) and 2 μg/mL doxycycline. For neuronal maintenance, half of the media was changed every 2-3 days. Cells were characterized for neuronal identity and maturation using immunocytochemistry and gene expression analysis (**Figure S1**). For immunocytochemistry, cells were fixed with 4% PFA for 15 min at room temperature, washed with DPBS containing Ca²⁺/Mg²⁺, permeabilized with 0.2% Triton X-100 for 15 min, and blocked for 1 h in DPBS containing 3% normal goat serum and 1% BSA. Cells were incubated overnight at 4 °C with anti-TUJ1 (1:1000; GTX85469, GeneTex) and anti-MAP2 (1:1000; ab32454, Abcam), followed by Alexa Fluor 647-conjugated secondary antibodies (1:500) for 1 h at room temperature. Nuclei were counterstained with DAPI. Images were acquired using a Leica DMIRB inverted microscope and processed in ImageJ.

### iPSC RNA-Seq processing

Cells for RNA-Seq were collected at four different timepoints during differentiation: day 1 of differentiation in vitro (DIV) or DIV1, DIV5, DIV9, and DIV14. RNA was extracted using the automated Promega Maxwell RSC instrument and the Maxwell RSC simplyRNA Tissue Kit (AS1340, Promega Corporation, Madison, Wisconsin, USA) according to the manufacturer’s recommendations.

rRNA-depleted RNA-Seq libraries from KOLF control and ReNU iPSC-derived neuronal differentiation samples were sequenced to a target depth of approximately 50 million reads per sample. RNA-seq data were processed using the nf-core/rnaseq pipeline (v3.22.2) executed with Nextflow v25.10.4 v2.7.11b. Read alignment and transcript quantification were performed using STAR and Salmon-based quantification modules on GENCODE release 49. RNA-seq quality metrics extracted from MultiQC included uniquely aligned reads, mapped read counts, duplication rate, proportion of proper pairs, 5′–3′ coverage bias and proportion of trimmed bases (**Figure S2A**).

Principal component analysis (PCA) was generated using the DESeq2 workflow implemented by nf-core/rnaseq. Briefly, transcript counts were normalized using DESeq2 and transformed using the variance-stabilizing transformation (VST). PCA coordinates were derived from the top 500 most variable genes identified across all samples using the pipeline default settings. PCA showed tight clustering of samples according to DIV stage and some separation of samples based on cell line (ReNU or KOLF; **Figure S2B**). Alternative splicing was quantified using rMATS-turbo (v4.3.0) against the GENCODE v49 annotation set with paired-end, unstranded settings (--libType fr-unstranded) and enabled novel splice-site detection (--novelSS).

## Results

### Consistent aberrant splicing events suggest an RNA-based biomarker for ReNU syndrome

We utilised two RNA-Seq datasets: (1) whole blood RNA-Seq samples from 11 ReNU patients and 33 controls (matched on age, sex, and sequencing depth) from the UK National Genomic Research Library (NGRL)^19^, and (2) lymphocyte RNA-Seq samples from 19 ReNU cases and 21 controls published in Nava *et al*.^3^ (**Figure 1A**).

For each cohort, we used rMATS to quantify alternative splicing events in ReNU cases and matched controls (see **Methods**). In total, 826,090 alternative splicing events were quantified in whole blood, and 826,806 in lymphocytes. Across both cohorts, 1,403,466 distinct events were observed, of which 249,430 (17.8%) were detected in both datasets. The remaining 1,154,036 events were quantified in only one cohort. Of these, 73.0% (842,029) involved one or more novel splice sites that were absent from gencode v49, indicating that most cohort-unique events were unannotated junctions not reproducibly detected across sample types.

We then assessed alternative splicing events found to be differentially spliced (FDR<0.05) between ReNU cases and matched controls in both the whole blood and lymphocyte cohorts. Of the 249,430 events quantified in both cohorts, 11,557 were differentially spliced in whole blood and 3,290 in lymphocytes, with 483 events significant in both cohorts (**Figure 1B**). PCA based on percent spliced in (PSI) values of the 483 shared events was dominated by PC1 (60.4%), reflecting differences in whole blood versus lymphocytes and/or underlying sample preparation and sequencing protocols rather than ReNU cases versus controls (**Figure 1C**).

Although base-line PSI values differed between sample types, we wondered if the magnitude of the ReNU-associated splicing shifts might nevertheless be conserved. We therefore compared the difference in PSI between ReNU cases and matched controls (Δlogit-PSI) separately in whole blood and lymphocytes across the 483 shared events. We found that effect sizes were not highly correlated for most event types (**Figure S3**), however were highly concordant among the 105 alternative 5’ splice site (A5SS) events (**Table S1**), consistent with the known biological role of U4 (Δlogit-PSI *R*^2^= 0.79). Of these events, 77/105 (73%) were not reported in the original analyses conducted on either cohort^1,3^. Of the 105 A5SS events, 95 had a concordant Δlogit-PSI direction between cohorts.

Among the 105 A5SS events that were significant in both cohorts, the change in splicing levels of any one splice-site was small: 90% had |ΔPSI| < 0.125 in whole blood and < 0.085 in lymphocytes. We therefore hypothesised that combining splicing effects across multiple ReNU-associated alternative splicing events would create a more robust splicing signature.

### Developing a six-event splicing signature for T-loop ReNU syndrome

To determine a minimum set of splicing events with high classification accuracy for ReNU syndrome, we randomly split 19 individuals (across both cohorts) with the n.64_65insT variant into training (N=14) and test (N=5) sets. We limited initially to only individuals with the n.64_65insT variant to avoid any differences associated with genotype. We filtered the 105 A5SS events that were significant in both cohorts to those with a minimum depth of 10 across all cases and controls, leaving a set of 84 candidate ReNU-associated A5SS events. We further limited our candidate space to one A5SS event per gene to avoid any overlapping or redundant sites, keeping the most discriminating site per gene. This left 77 candidate A5SS events (**Figure S4**).

We ran L1-penalised binary logistic regression on case/control status among the 14 training cases and all 54 controls, using Δlogit-PSI values for the 77 candidate A5SS events as features, adjusted so that all ReNU-associated splicing deviations were positive. We used leave-one-out cross validation with one fold for each of the 14 cases, where in each fold one case and a rotating subset of 3-4 controls were left out. This approach identified a minimal six-splice-site signature (**Figure 2A, Table S2**) that distinguished cases from controls with cross-validation AUC=0.93 (bootstrap 95% CI: 0.79-1.00). The final model perfectly separated cases from controls, including the five hold-out cases (two whole blood + three lymphocytes; AUC=1.00; **Figure 2B**). To check for overfitting, we repeated this process with 14 training samples and 5 test samples randomly selected from the 5,984 whole blood controls. Over 1,020 permutations the median training AUC was 0.68.

**Figure 2:**
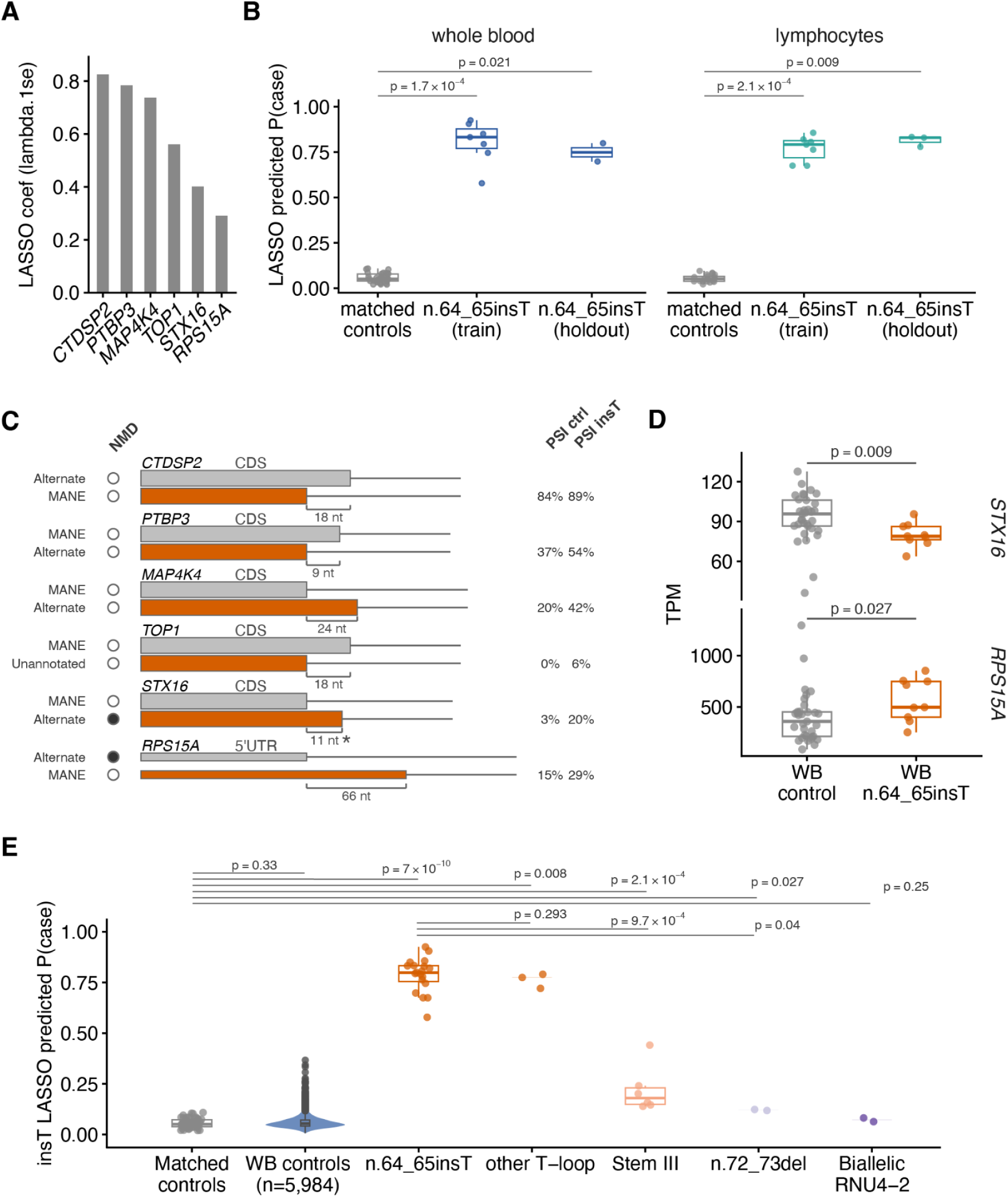
Development of a six-event T-Loop splicing signature. **A)** LASSO coefficients for the six A5SS events selected by the LASSO classifier at λ_1se_. Events are labelled by associated gene symbol, see **Table S2** for full event details. **B)** Predicted probability of case status generated by the LASSO-derived splicing signature, across whole blood and lymphocyte controls, training and holdout cases. Predictions were generated using the final λ_1se_ model. Mann-Whitney P-values are shown for training and holdout versus matched controls. Boxes represent the interquartile range with median. For holdout groups only individual points and median line are shown due to small sample size. **C)** Schematic of the six events comprising the n.64_65insT splicing signature. Grey and orange transcripts indicate decreased and increased 5′ splice-site usage, respectively, in n.64_65insT individuals. Transcripts are annotated as MANE Select, alternate, or unannotated; filled circles indicate predicted nonsense-mediated decay (NMD). Brackets indicate the distance (nt) between alternative 5′ splice sites, and the asterisk denotes a frameshift. Median PSI values in whole blood controls and n.64_65insT individuals are shown for the isoform with increased usage (orange). **D)** *STX16* and *RPS15A* TPM, in whole blood, quantified with Salmon, from n.64_65insT individuals and controls. Significance was assessed using two-sided Mann–Whitney U tests with Benjamini-Hochberg correction across the two genes. **E)** Application of the six-site signature to 5,984 unmatched whole-blood RNA-seq controls, matched controls used in signature development, and all available *RNU4-2* genotype groups. For unmatched controls, only outliers (n=211) are plotted as individual points because of sample size. Boxes represent the interquartile range with median. For groups with n ≤ 3 only individual plots and median lines are plotted. *P* values indicate comparisons with matched controls (top) and with n.64_65insT individuals (bottom) using two-sided Mann–Whitney U tests with Benjamini-Hochberg correction.

The six A5SS events in the final model impacted *CTDSP2, PTBP3, MAP4K4, TOP1, STX16* and *RPS15A* (**Figure 2C**). Five events were shifts in usage between nearby annotated 5’SS: *PTBP3*, *MAP4K4* and *STX15* shifted towards usage of an alternative transcript 5’SS and away from the MANE Select transcript 5’SS, whereas *CTDSP2* and *RPS15* were instead shifted towards usage of the MANE Select transcript 5’SS. The remaining event involved increased usage of an unannotated cryptic 5’SS in *TOP1*. Five of the six events impact coding sequences, with four of these predicted to preserve the reading frame. The exception is the event in *STX16* which introduces a frameshift and increased usage of an alternative transcript annotated as a nonsense mediated decay transcript (ENST00000460655.5).

The median PSI of the alternative splice-site increased from 3.0% in controls (PSI range 0.6-7.3%) to 19.5% in n.64_65insT individuals (PSI range 9.6-22.2%), with a ΔPSI of +16.5%. This was accompanied by a shift in TPM from a median of 95.7 in controls (TPM range 36.2-127.6) to 78.9 in n.64_65insT individuals (TPM range 63.8-95.5), consistent with the loss of transcripts through nonsense-mediated decay. The sixth event occurred within the 5’UTR of *RPS15A* and did not alter the coding sequence, but shifted splicing away from a 5’ splice site used only in nonsense mediated decay transcripts (ENST00000562935.5, ENST00000569365.6, ENST00000572008.5). *RPS15A* expression was significantly increased in n.64_65insT individuals (Mann-Whitney P = 0.027, **Figure 2D**). The median PSI of the alternative splice-site increased from 14.8% in controls (PSI range 8.1-28.8%) to 29.1% in n.64_65insT individuals (PSI range 22.4-42.9%), with a ΔPSI of +14.3%. This was accompanied by a shift in TPM from a median of 358.9 in controls (TPM range 87.3-1294.6) to 497.4 in n.64_65insT individuals (TPM range 253.4-853.2). None of the remaining genes showed a significant change in TPM in n.64_65insT individuals (**Figure S5A**).

We further assessed the specificity of our splicing signature using 5,984 whole blood samples available in the NGRL that were not selected as matched controls for splicing signature development. Of the six splicing signature events, five could be quantified in 100% of samples. The A5SS event in *TOP1* could not be quantified in 317/5,984 (5.3%) owing to neither splice junction having any supporting reads. PSIs for each site in whole blood were not significantly different between the set of 5,984 controls and the 33 controls used for splicing signature development for any of the six signature events, whereas the ReNU cases were significantly different to the 5,984 controls for all six sites (**Figure S6A**). Applying the LASSO model, probabilities calculated on the 5,984 unmatched controls did not significantly differ from the matched controls (p = 0.33, **Figure 2E**). The highest scoring unmatched control was 0.37, which was below the lowest scoring n.64_65insT individual (0.58). PCA of the splicing signature across all 5,984 individuals showed no clustering by age, sex or sequencing depth (**Figure S6B**). We also noted that an additional whole blood RNA-Seq sample from an individual with the n.64_65insT variant, which had been excluded from model development due to being an extreme outlier in total reads, received a score of 0.69 using the splicing signature.

Using any one of the signature events individually results in between 21 and 5,664 false positives in the wider control cohorts, defined as a control sample having a larger weighted Δlogit-PSI than the lowest scoring n.64_65insT individual (**Figure S6C**). We assessed how many sites were required to achieve high specificity by taking all possible combinations of two to five signature sites and constructing scores from summing their weighted Δlogit-PSI across the selected events. Combining multiple events progressively improved specificity, with some combinations of three or more events capable of achieving zero false positives (**Figure S6C**).

Having established a six-event n.64_65insT-associated splicing signature, we next assessed whether this signature generalised to other *RNU4-2* genotypes. When applied to the remaining ReNU cases, the three individuals with other T-loop variants were also assigned high probabilities (median = 0.775, **Figure 2E**) significantly greater than matched controls (Mann-Whitney P = 0.008, median = 0.051) and not significantly different to individuals with n.64_65insT (Mann-Whitney P = 0.293, median = 0.799). In contrast, the six individuals with variants in Stem III and the two individuals with the n.72_73del variant that falls between the T-loop and Stem III had much lower probabilities (median = 0.179 and 0.121 respectively; **Figure 2E**), far closer to, but still significantly greater than controls (Mann-Whitney P = 2.1×10^-4^; P = 0.027). Two individuals with biallelic variants in *RNU4-2* leading to recessive *RNU4-2* associated NDD^10^ likewise had low predicted probabilities (median = 0.072, **Figure 2E**). Collectively, these results indicate that the signature is specific to the dominant T-loop-associated form of ReNU syndrome rather than *RNU4-2* dysfunction more broadly.

### Stem III variants exhibit a partially overlapping but distinct splicing signature

Motivated by the distinct splicing profile observed in individuals with T-loop variants, we explored whether there were distinct Stem III-specific splicing changes. Owing to the limited number of Stem III RNA-seq samples (n = 6), we were not able to create a robust Stem III splicing signature with hold-out samples. Nevertheless, we used the same classifier approach to attempt to identify Stem III specific events using all available samples for model training and cross-validation (**Figure S4**). L1-penalised logistic regression resulted in a model made up of six A5SS events, with two events shared with the T-loop splicing signature (**Figure 3A, Table S3**). This included an event in *MAP4K4* that was previously noted to be shared between T-loop and Stem III individuals^3^ and the event in *TOP1*.

**Figure 3:**
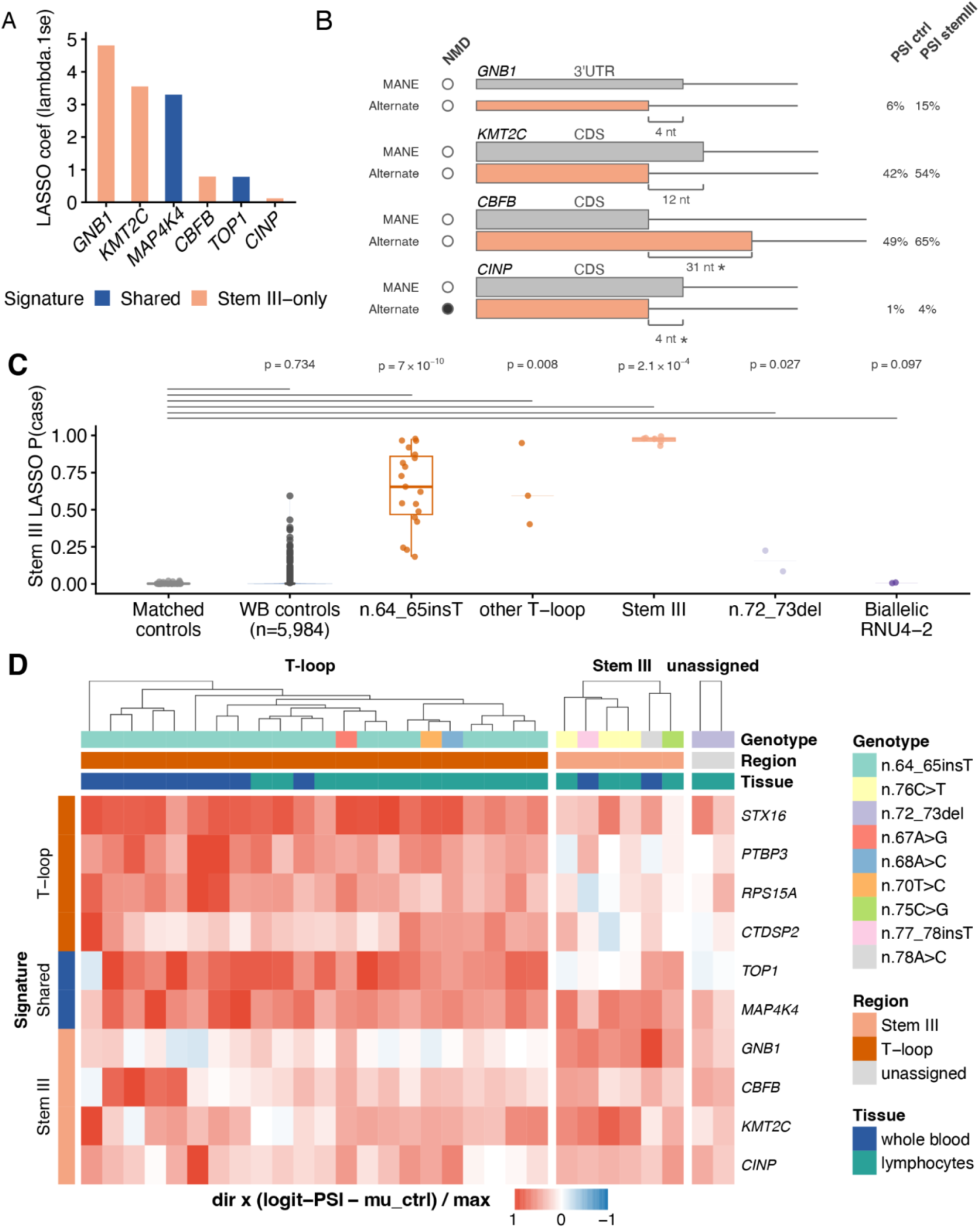
Development of a Stem III splicing signature. **A)** LASSO coefficients for the six A5SS events selected by the LASSO classifier at λ_1se_. Events are labelled by associated gene symbol, and coloured blue if they are shared with the T-loop splicing signature. **B)** Schematic of the four Stem III-only events. Grey and orange transcripts indicate decreased and increased 5′ splice-site usage, respectively, in Stem III individuals. Transcripts are annotated as MANE Select, alternate, or unannotated; filled circles indicate predicted nonsense-mediated decay (NMD). Brackets indicate the distance (nt) between alternative 5′ splice sites, and the asterisk denotes a frameshift. Median PSI values in whole blood controls and Stem III individuals are shown for the isoform with increased usage (orange). **C)** application of the Stem III signature to 5,984 unmatched whole-blood RNA-seq controls, matched controls used in signature development, and all available *RNU4-2* genotype groups. For unmatched controls, only outliers are shown because of sample size. Boxes represent the interquartile range with median. *P* values indicate comparisons with matched controls (top) using two-sided Mann–Whitney U tests with Benjamini-Hochberg correction. **D)** Heatmap of splicing perturbation across all T-loop and Stem III model events in ReNU cases. Cell values represent delta logit PSI values per site normalised so that ReNU associated shifts are positive (red). Values were additionally divided by the maximum values for that event across all ReNU samples. Rows are grouped into T-loop signature events (top), events shared between the T-loop and Stem III splicing signatures (middle), and Stem III-specific events (bottom). Columns are grouped by variant region and annotated by genotype and tissue source.

The four A5SS events included only in the Stem III event set affected *GNB1*, *KMT2C*, *CBFB* and *CINP* (**Figure 3B**). All four events represented shifts in usage away from the MANE Select 5′ splice site towards an annotated alternative splice site. Two events (*CBFB* and *CINP*) induced frameshifts: the *CINP* event increased usage of an alternative transcript annotated as undergoing nonsense-mediated decay, whereas the *CBFB* event introduced a frameshift close to the 3′ end of the coding sequence and corresponds to an annotated alternative protein-coding transcript in Ensembl. Given only two Stem III whole-blood samples were available, we could not reliably test for differences in transcript abundance in Stem III variant carriers compared with controls (**Figure S5B**).

We next assessed whether the six A5SS events identified using the Stem III samples could predict other *RNU4-2* genotypes. Individuals with n.64_65insT and other T-loop variants had elevated predicted probabilities (median = 0.654 and 0.594, respectively), which were significantly greater than matched controls (median = 0.001; *P* = 7 x 10^-10^ and *P* = 0.008 for n.64_65insT and other T-loop, respectively). Individuals with the n.72_73del variant, which falls between the T-loop and Stem III, had predicted probabilities that were significantly higher than controls, but that were still relatively low (median = 0.155; *P* = 0.027 versus matched controls). The two individuals with the recessive *RNU4-2* condition also had low predicted probabilities (median = 0.007) that were comparable to controls (*P* = 0.097).

To better understand the genotype specificity of the T-loop and Stem III A5SS events, we examined the per-event splicing deviations across all ReNU samples (**Figure 3D**). The highest-weighted event in the Stem III model, *GNB1*, was largely restricted to individuals with Stem III variants. In contrast, the remaining three Stem III-model events (*KMT2C*, *CBFB* and *CINP*) showed variable perturbation across Individuals with both Stem III and T-loop variants. Thus, although these splicing changes appear stronger in individuals with Stem III variants, several constituent events were shared with other dominant *RNU4-2* genotypes.

The splice sites selected in the classifiers for both T-loop and Stem III variants were consistent with previously reported features of ReNU-associated alternative splicing^3^. Specifically, donor splice sites showing increased usage had lower predicted splice strengths than those with decreased usage (**Figure S7A**), and sequence logos recapitulated the characteristic depletion of AAG at positions +3 to +5 in splice sites with reduced usage, accompanied by increased reliance on A/G nucleotides at positions −2 and −1 among splice sites with increased usage (**Figure S7B**).

### The T-loop splicing signature generalises to iPSCs

We next examined whether ReNU-associated splicing abnormalities identified in blood-derived tissues were recapitulated in a patient-derived neuronal iPSC model. RNA-seq was generated from three clones derived from the same patient harbouring the 64_65insT variant at four stages of neuronal differentiation (see **methods**). Alternative splicing events were quantified using rMATS and compared to KOLF control cells (see **methods**).

Across the four stages of neuronal differentiation, 45,162 unique alternative splicing events were differentially used between ReNU and KOLF cells in at least one stage, with skipped exons comprising the largest event class (**Figure S8A**). Of these events, 5,727 (12.6%) were significantly different in more than one stage of differentiation with 4,702 showing concordant direction of splicing change in all stages where they were significant (significantly more than expected by chance, Poisson-binomial P = 3.3×10^-16^). A5SS events were most likely to be shared and directionally concordant across multiple timepoints when compared with all other event types (**Figure S8B,** OR = 1.32, Fisher’s exact P = 1.9×10^-19^).

We next asked whether the 95 A5SS events that showed concordant ReNU-associated splicing changes in the whole-blood and lymphocyte cohorts (**Table S1**) were recapitulated in iPSCs. 78 of the 95 events (82%) could be quantified across all four stages of neuronal differentiation. At individual differentiation stages, 82–83 of the 95 candidate events could be quantified, of which 23–36 (28.0–43.4%) were significantly altered between ReNU and KOLF cells at FDR < 0.05 (**Figure 4A**, **Table S4**). This was substantially greater than the 3.2-5.8% expected among randomly selected A5SS events quantified in all three datasets, representing a 7.3-8.7-fold enrichment (hypergeometric *P* = 1.24×10^-15^ to 2.05×10^-24^; **Figure 4A**). Furthermore, Δlogit-PSI values were remarkably conserved across whole blood, lymphocytes, and iPSCs (R² = 0.59–0.75 for all events; R² = 0.85–0.89 for events at FDR < 0.05 in iPSCs; **Figure 4B**).

**Figure 4:**
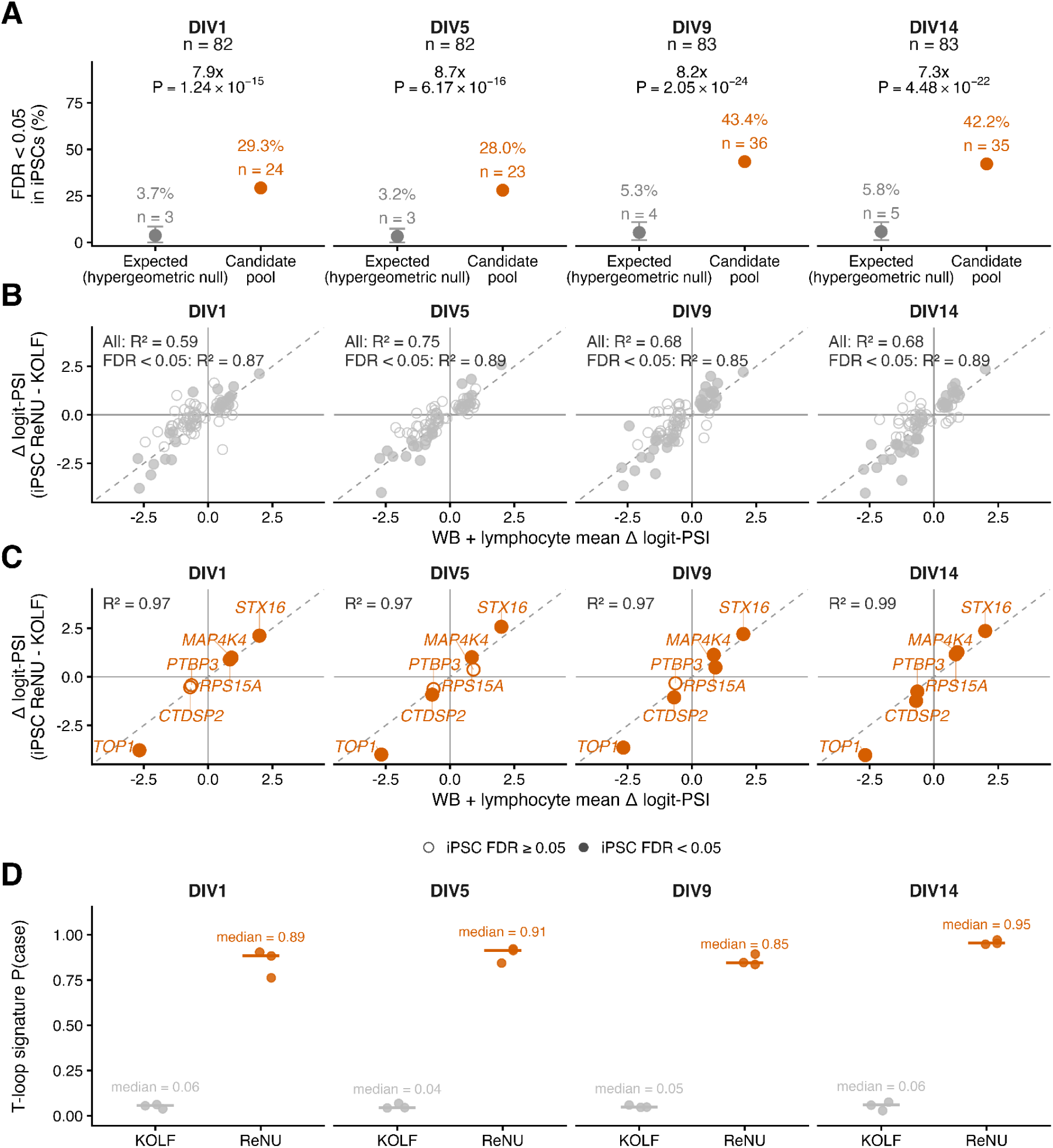
Blood-derived ReNU-associated splicing events and the T-loop signature are recapitulated in iPSC-derived neurons. **A)** Enrichment of ReNU-associated A5SS candidate events among events significantly altered in iPSC-derived neurons (FDR < 0.05). At each differentiation stage (DIV1, DIV5, DIV9 and DIV14), 82, 82, 83 and 83 candidate events, respectively, were quantifiable in iPSCs. Orange points show the proportion and number of quantifiable candidate events reaching FDR < 0.05 between ReNU and KOLF cells. Grey points show the proportion expected under a hypergeometric null, defined using A5SS events quantifiable in whole blood, lymphocytes and the corresponding iPSC differentiation stage. error bars indicate the central 95% quantile interval of the hypergeometric null distribution for a random sample of the same size as the quantifiable candidate set. Fold enrichment and exact upper-tail hypergeometric *P* values are shown above each comparison. **B)** Correlation of Δlogit-PSI values between the iPSC model and the mean Δlogit-PSI across individuals with n.64_65insT across whole blood and lymphocyte cohorts for the A5SS candidate pool. Open and filled points indicate events with iPSC FDR ≥ 0.05 and FDR < 0.05, respectively. R² values are shown for all quantifiable candidate events and separately for the subset reaching FDR < 0.05 in iPSCs. **C)** Correlation of Δlogit-PSI values for the six A5SS events comprising the T-loop splicing signature. Axes and point fills are defined as in **(B)**; signature events are shown in orange and labelled by gene. **D)** Predicted probability of case status generated by the T-loop splicing signature for ReNU and KOLF iPSC samples at each stage of neuronal differentiation. Each point represents an independent iPSC clone. horizontal lines indicate group medians, with median probabilities annotated.

Restricting the analysis to the six A5SS events comprising the final T-loop splicing signature, all six showed the same direction of ReNU-associated splicing change in the neuronal and blood-derived datasets at every differentiation stage. The events in *STX16*, *RPS15A* and *TOP1* reached FDR < 0.05 at all four stages, and all six signature events were FDR-significant by DIV14. Irrespective of whether individual events reached FDR significance at each stage, effect sizes for the six signature events showed near-perfect correlations between the neuronal and blood-derived datasets across all four differentiation stages (R² = 0.97–0.99; **Figure 4C**).

Finally, we applied the six-site T-loop splicing signature, trained exclusively using blood-derived data, to the ReNU iPSC model across all four stages of neuronal differentiation. The signature consistently assigned high probabilities of ReNU status to ReNU cells (median predicted probabilities 0.89, 0.91, 0.85, and 0.95 at DIV1, DIV5, DIV9 and DIV14, respectively) and low probabilities to KOLF controls (median predicted probabilities 0.06, 0.04, 0.05, and 0.06 at DIV1, DIV5, DIV9 and DIV14, respectively) across all four stages of neuronal differentiation (**Figure 4D**).

## Discussion

Here we define a six-site RNA splicing signature for T-loop ReNU syndrome from alternative 5′ splice-site (A5SS) events detected in RNA-Seq data from two patient cohorts. The signature is robust across sequencing methods, tissues, and *in vitro* model systems, is specific to variants in the U4 T-loop, and perfectly discriminates between cases and controls. Together, these findings establish a molecular biomarker of splicing dysregulation in ReNU syndrome with immediate relevance to therapeutic development for ReNU syndrome.

The splicing signature shows high accuracy across sample types. Comparing alternative splicing across cohorts is often challenging due to batch effects and tissue-specific splice-site usage^8,9^. Nevertheless, we identified consistent A5SS events associated with ReNU syndrome in RNA-Seq data from whole blood, cultured lymphocytes, and patient iPSC-derived neurons. Importantly, the selected A5SS events had similar effect sizes between tissues despite differences in baseline PSI. By using Δlogit-PSI values, we avoid compression of effect sizes for PSI values approaching zero or one^12,13,21^, allowing quantitative comparisons across distinct contexts and enhancing the portability of the splicing signature. Although the signature generalised remarkably well to a patient iPSC-derived neuronal model, validation was limited to a single iPSC lineage. Additional patient-derived or CRISPR-engineered models, with isogenic controls, will be required to establish robustness across genetic backgrounds.

The individual events selected for the splicing signature were subtle shifts in splice-site usage consistent with ReNU syndrome variants causing global but low level disruption to splicing. Most events were shifts in usage at annotated splice sites outside of known NDD genes that were not predicted to result in NMD. The selected events are unlikely to be the principal drivers of ReNU syndrome pathophysiology, but instead represent splicing changes which are both tolerated at a high enough level to reach statistical significance and that are highly represented in RNA-Seq reads. More damaging events which disrupt transcript stability may be challenging to quantify, and therefore have limited utility as biomarkers. The preference for nearby alternative donor sites which are also used in control samples is consistent with previous findings, suggesting that altered specificity of 5′ splice-site recognition disrupts competition between known 5’SS rather than broadly activating cryptic splice sites^3^.

Our findings provide evidence that variants in the T-loop and Stem III have partially distinct molecular consequences. The T-loop signature, developed using individuals with the recurrent n.64_65insT variant, did not generalise to Stem III individuals. Instead, we found preliminary evidence for Stem III-specific events, most notably an A5SS event in *GNB1*. That Stem III variants should evince a distinct molecular phenotype is consistent with a recent saturation genome editing experiment in which variants in the T-loop, but not those in Stem III, showed an effect in a diploid cell line^5^. These data are consistent with a background splicing defect common to all ReNU syndrome variants, overlaid with variant-specific perturbation of particular splice sites. Interestingly, individuals with the n.72_73del variant, which is located between the T-loop and Stem III, did not have high predicted probabilities with either the T-loop or Stem III models. Only having two samples with this specific genotype, however, prevented us from investigating this further. Larger cohorts will enable more complete characterisation of these distinct molecular phenotypes.

The splicing signature is a continuous, quantitative measure of splicing dysfunction that is reproducible across tissues and experimental systems and which reflects underlying disease biology. We therefore envision that it will have significant utility as a quantitative biomarker for therapeutic development, particularly as a candidate pharmacodynamic biomarker under the BEST (Biomarkers, EndpointS and other Tools) framework^22^. The biomarker could have utility in both pre-clinical and clinical therapeutic design, development, and monitoring, as a direct assay of target engagement and treatment response. In principle, targeted sequencing or even multiplexed quantitative PCR of the selected loci would suffice for high-throughput signature quantification. *In vitro*, the splicing signature bridges the gap between molecular proxies of treatment effect (such as allele-specific RNA abundance) and cellular read-outs (such as viability or cellular phenotype). *In vivo*, the splicing signature may enable an early and objective measure of treatment response. Validation in clinically relevant samples, especially cerebrospinal fluid, will be important for future therapeutic agents delivered intrathecally. Despite the potential high utility of the splicing signature, we note that the quantitative relationship between mutant *RNU4-2* dosage, the magnitude of splicing correction and clinical phenotype remains unknown. Correlating the splicing signature with therapeutic benefit could ultimately enable its use as a surrogate endpoint for therapeutic efficacy.

In summary, we describe a quantitative, RNA-based splicing signature for T-loop ReNU syndrome. The signature has immediate significance for therapeutic development in this prevalent disorder. More broadly, disruption to splicing is a common feature of spliceosomopathies; this work offers a framework for RNA biomarker discovery to accelerate therapy development across these disorders.

## Data Availability

Data from the National Genomic Research Library (NGRL) used in this research are available within the secure Genomics England Research Environment (GERE). Access to NGRL data is restricted to adhere to consent requirements and protect participant privacy. Data used in this research included whole blood RNA-seq data generated for the 100kGP transcriptomics pilot and extension. More details, including paths to these data within the GERE, are available here: https://re-docs.genomicsengland.co.uk/rna_seq/. Access to NGRL data is provided to approved researchers who are members of the Genomics England Research Network, subject to institutional access agreements and research project approval under participant-led governance. For more information on data access, visit: https://www.genomicsengland.co.uk/research
RNA-seq data for the lymphocyte cohort are available through the European Genome-phenome Archive (EGA) under accession EGAS50000000889 (https://ega-archive.org/studies/EGAS50000000889). Raw RNA-seq data from patient-derived iPSCs cannot be shared given privacy and consent regulations under which the original fibroblasts were obtained during routine clinical diagnostics.
Code used for data processing, statistical analyses, development of the splicing signature, and generation of the figures is available at https://github.com/Computational-Rare-Disease-Genomics-WHG/rnu42_splicing_biomarker.

https://www.genomicsengland.co.uk/research

https://ega-archive.org/studies/EGAS50000000889

https://github.com/Computational-Rare-Disease-Genomics-WHG/rnu42_splicing_biomarker

## Acknowledgements

We sincerely thank the patients and their families who generously donated data and samples, without which this work would not have been possible.

This work was funded by the Medical Research Council Centre of Research Excellence in Therapeutic Genomics (grant number MR/Z504725/1 to C.R., S.J.S. and N.W.), the Health Data Research UK QQ2 Molecules to Health Records Driver Programme (to S.J.S), the National Institute of Mental Health (grant number R01MH129751 to S.J.S.), a Wellcome Career Development Award (grant number 305292/Z/23/Z, to N.W.) and a Lister Institute research prize (to N.W.). R.D. is supported by a National Health and Medical Research Council (NHMRC) Investigator Grant (Emerging Leadership 1, grant number 2041062). The Barakat lab was supported by the Netherlands Organisation for Scientific Research (ZonMw Vidi, grant number 09150172110002), and a research grant (WAR26-18) for ReNU syndrome from the Sophia Research Foundation (Stichting Sophia Kinderziekenhuis Fonds) and acknowledges other ongoing and former support for rare disease research from Stichting 12q, EpilepsieNL, CURE Epilepsy, Spastic Paraplegia Foundation, Inc. Funding bodies did not have any influence on study design, results, and data interpretation or final manuscript.

We gratefully acknowledge the participants of the National Genomic Research Library (NGRL), whose contributions made this research possible. Secure access to the NGRL under project ID RR354 was provided by Genomics England, which delivers the NGRL in partnership with NHS England, and is wholly owned by the UK Department of Health and Social Care. The NGRL contains participants’ health data collected by the NHS as part of their care, along with samples and data from their participation in research, for which fully informed consent has been obtained. This includes genomic and clinical data provided through the NHS Genomic Medicine Service, as well as data obtained through research studies, including the 100,000 Genomes Project and the Generation Study, both of which are delivered in partnership with the NHS, and from other research cohorts involving external collaborators.

## Author Contributions

R.D., A.B., S.J.S. and N.W conceived the study. R.D., A.B., and M.G. performed data analysis. R.D. created the figures. T.M. performed experiments in iPSCs. B.C., T.S.B., and C.R. provided materials and reagents. J.C.B. provided guidance on statistical analysis. S.J.S. and N.W. supervised and funded the work. All authors reviewed and commented on the final manuscript.

## Data and Code availability

Data from the National Genomic Research Library (NGRL) used in this research are available within the secure Genomics England Research Environment (GERE). Access to NGRL data is restricted to adhere to consent requirements and protect participant privacy. Data used in this research included whole blood RNA-seq data generated for the 100kGP transcriptomics pilot and extension. More details, including paths to these data within the GERE, are available here: https://re-docs.genomicsengland.co.uk/rna_seq/. Access to NGRL data is provided to approved researchers who are members of the Genomics England Research Network, subject to institutional access agreements and research project approval under participant-led governance. For more information on data access, visit: https://www.genomicsengland.co.uk/research

RNA-seq data for the lymphocyte cohort are available through the European Genome-phenome Archive (EGA) under accession EGAS50000000889 (https://ega-archive.org/studies/EGAS50000000889). Raw RNA-seq data from patient-derived iPSCs cannot be shared given privacy and consent regulations under which the original fibroblasts were obtained during routine clinical diagnostics.

Code used for data processing, statistical analyses, development of the splicing signature, and generation of the figures is available at https://github.com/Computational-Rare-Disease-Genomics-WHG/rnu42_splicing_biomarker.

## Supplementary Figures

**Supplementary Figure 1:**
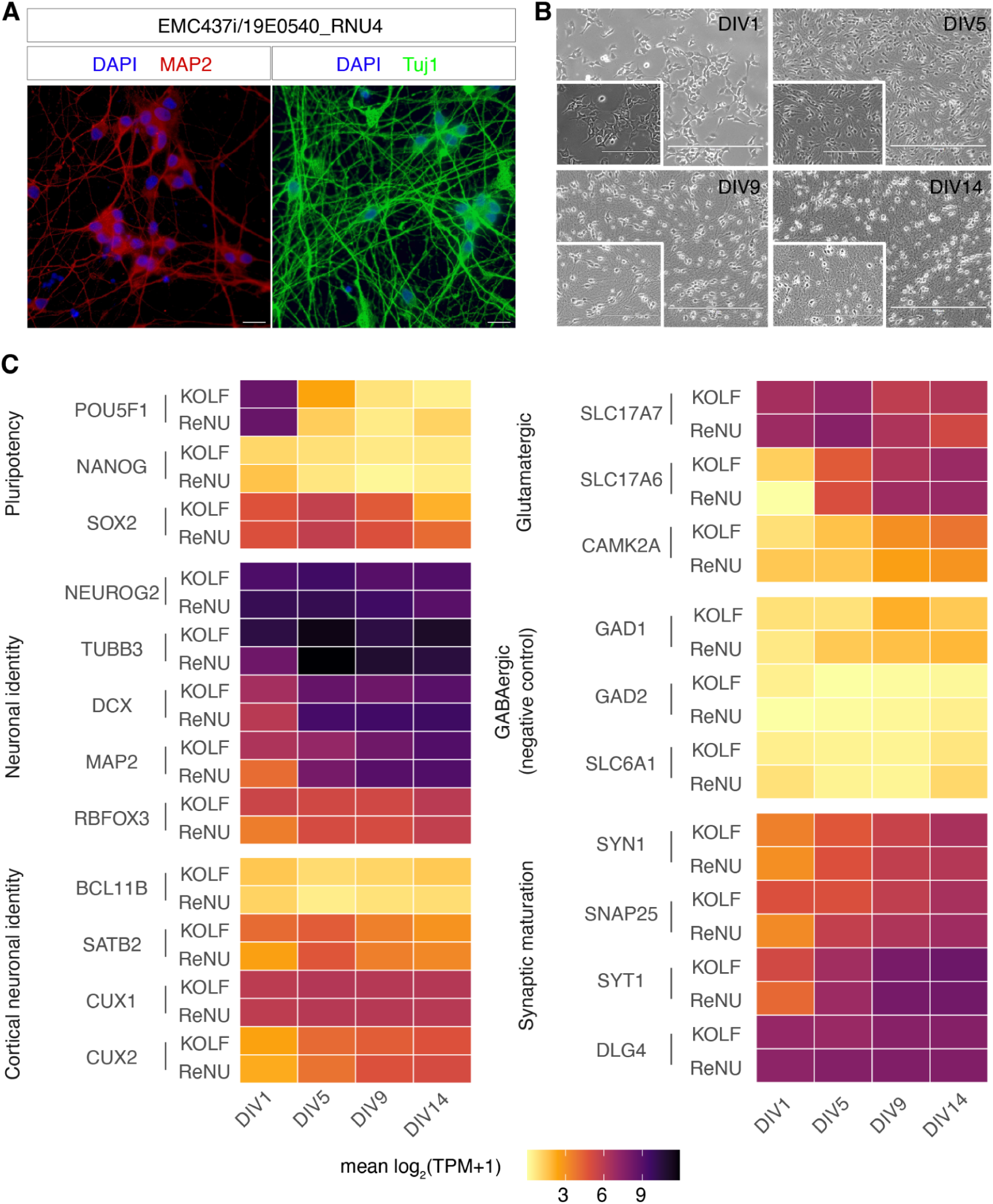
Characterization of iPSC-derived neuronal differentiation and maturation. **A)** DIV14 iPSC-derived neurons stained for neuronal markers MAP2 and TUJ1, along with DAPI for nuclear staining. Expression of MAP2 and TUJ1, along with extensive neurite networks, confirms successful differentiation into neurons. Scale bars: 20 µm. **B)** Brightfield microscopy images depicting the dynamic changes in cell morphology at the collection timepoints (DIV1, DIV5, DIV9, and DIV14). Insets of increased focus (20X) highlight regions of interest, emphasizing neurite outgrowth and cell aggregation as key features of differentiation. Scale bars: 400 µm (for 10X images); insets (20X): 200 µm **C)** TPM of marker genes across differentiation timepoints, quantified with Salmon. Mean log2(TOM + 1) values were calculated across the control line replicates (KOLF, n = 3) and the *RNU4-2* n.64_65insT replicates (ReNU, n = 3) within each timepoint.

**Supplementary Figure 2:**
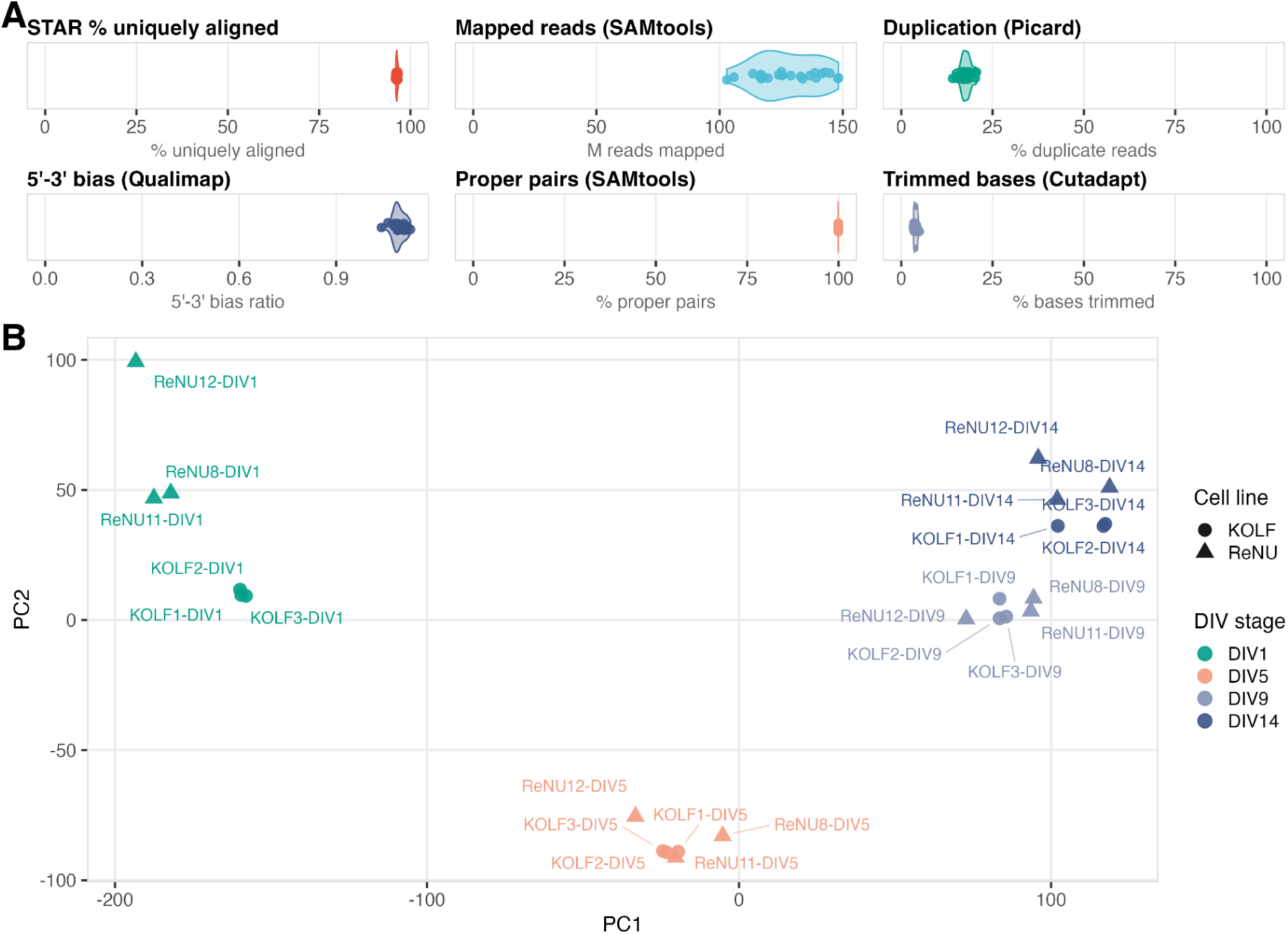
Quality-control metrics of patient-derived iPSC neuronal models. **A)** Distribution of per-sample RNA-seq quality-control metrics across the 24 iPSC-derived neuronal differentiation samples (n=3 clones derived from the same patient harbouring the mutation 64_65insT compared with n=3 KOLF controls, each with 4 stages of differentiation in-vitro). For Trimmed bases values from Cutadapt, values from paired fastq files were averaged to generate a single per-sample estimate before plotting. **B)** Principal component analysis of transcriptome profiles generated by the nf-core/rnaseq DESeq2 workflow. Principal components were calculated after DESeq2 normalization and variance-stabilizing transformation (VST) of transcript counts using the standard nf-core/rnaseq DESeq2 analysis workflow. The plotted PCA was generated from the top 500 most variable genes across all samples. Points are coloured according to differentiation stage (DIV1, DIV5, DIV9 and DIV14) and shaped according to cell line (KOLF, circle or ReNU, triangle).

**Supplementary Figure 3.**
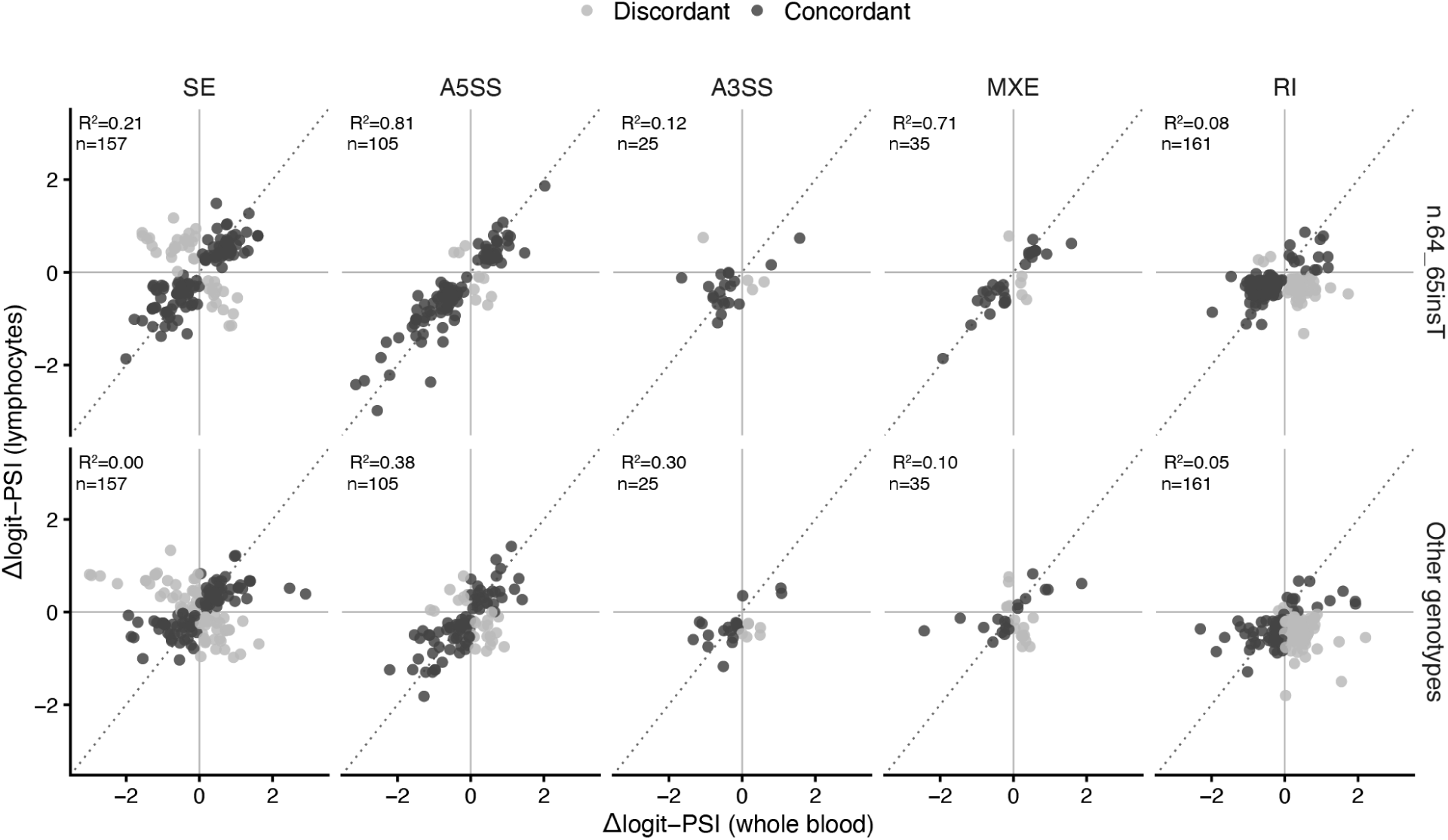
Consistency of alternative splice site events in whole blood and lymphocyte cohorts. Scatterplots show the mean Δlogit-PSI values across ReNU cases versus controls in whole blood versus Lymphocyte datasets for 483 alternative splicing events significant in both cohorts. Panels are split into splicing event classes as annotated by rMATS: skipped exons (SE), alternative 5′ splice sites (A5SS), alternative 3′ splice sites (A3SS), mutually exclusive exons (MXE) and retained introns (RI). Points are coloured by whether the effect direction is concordant (dark grey) or discordant (light grey) between cohorts. R² and event counts are shown for each plot.

**Supplementary Figure 4.**
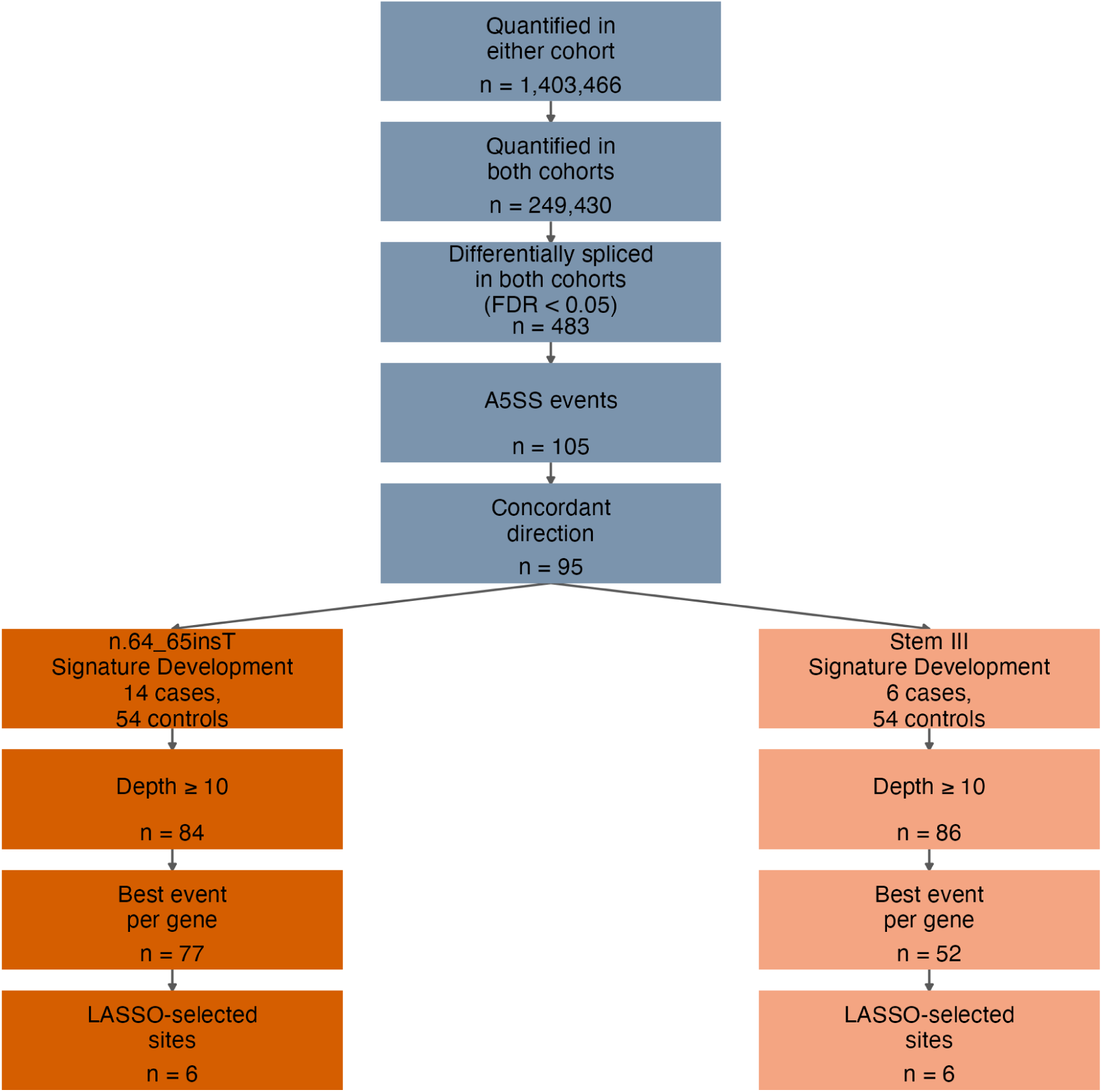
Filtering of alternative splicing events for signature development. A total of 1,403,466 alternative splicing events were quantified by rMATS-turbo^20^ in at least one of the whole blood and lymphocyte cohorts, of which 249,430 were quantified in both cohorts. Statistical filtering (FDR < 0.05) identified 483 events significant in both, including 105 A5SS events, 95 of which had a concordant direction of Δlogit-PSI between the two cohorts. For signature development, events were filtered separately for the n.64_65insT and Stem III signatures based on minimum read depth across all cases and controls used in training (≥10 reads; 84 and 86 events, respectively), followed by selection of the best event per gene as defined by the geometric mean of a Mann-Whitney AUC across cohorts (77 and 52 events, respectively). Least absolute shrinkage and selection operator (LASSO) regression identified six A5SS events for the final insT signature and six A5SS events for the final Stem III signature.

**Supplementary Figure 5.**
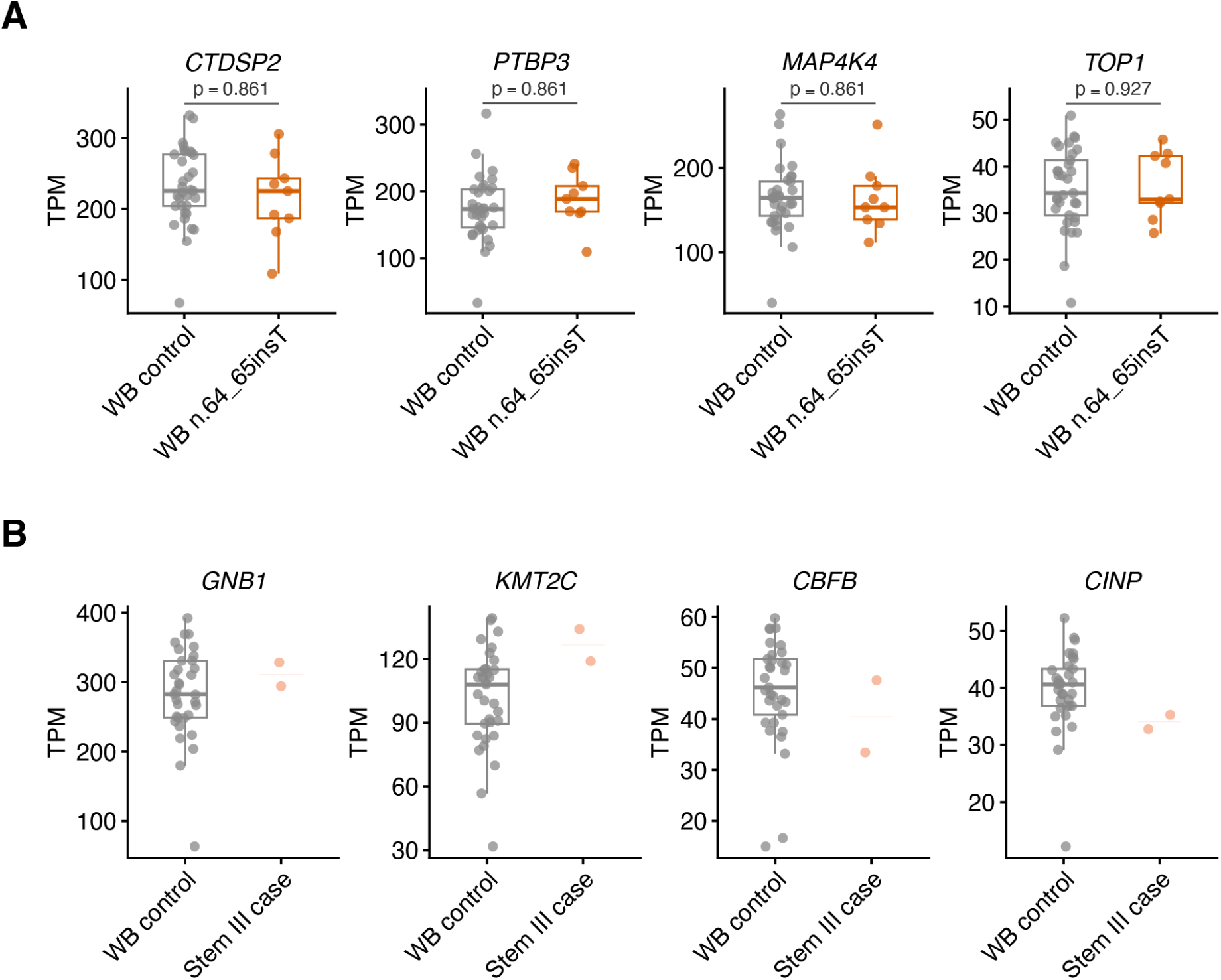
**A)** *CTDSP2, PTBP3, MAP4K4* and *TOP1* TPM in whole blood from n.64_65insT individuals and controls, quantified with Salmon. Significance was assessed using two-sided Mann–Whitney U tests with Benjamini-Hochberg correction across the four genes. Boxplots show medians and interquartile ranges (IQRs) across combinations of that number of sites; whiskers extend to the most extreme points within 1.5 × IQR. **B)** *GNB1, KMT2C, CBFB* and *CINP* TPM in whole blood from Stem III individuals and controls, quantified with Salmon. Boxplots show medians and interquartile ranges (IQRs) across combinations of that number of sites; whiskers extend to the most extreme points within 1.5 × IQR. For Stem III cases only individual points with a median line are shown due to small sample size.

**Supplementary Figure 6:**
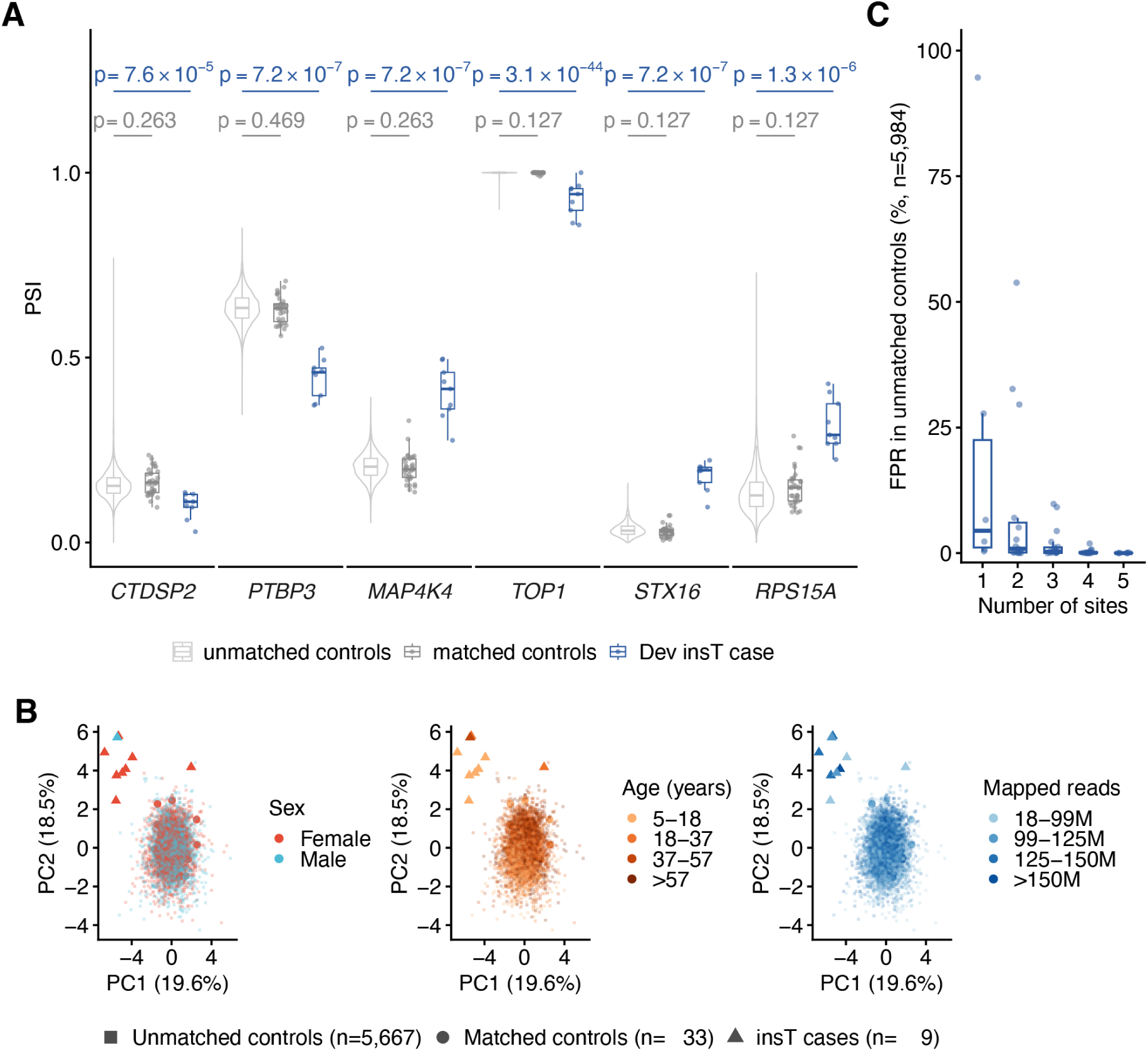
Validation and Specificity of the ReNU splicing signature in 5,984 whole blood RNA-Seq samples from the Genomics England 100kGP Transcripts Pilot and Extension. **A)** Per-site PSI distributions for the six splice events retained in the final λ_1se_ LASSO classifier. Three groups are shown for each event: 5,984 rare disease probands in the NGRL (‘unmatched controls’; light grey), 33 controls matched with whole blood ReNU cases (‘matched controls’; dark grey) and nine whole blood ReNU cases with the n.64_65insT variant (‘Dev insT case’; blue). The unmatched cohort is displayed as a boxplot with violin density distribution, whereas development-cohort samples are shown as boxplots with horizontally jittered points. Boxplots show medians and interquartile ranges (IQRs); whiskers extend to the most extreme points within 1.5 × IQR. Two-sided Mann–Whitney U-tests were performed for each event comparing development controls versus unmatched controls and development ReNU cases versus unmatched controls. P values for the 12 tests were adjusted jointly using the Benjamini–Hochberg procedure. **B)** PCA of logit PSI values for the six selected splicing signature events among the whole blood cohort. Unmatched controls (small squares), matched controls (large circles) and n.64_65insT cases (large triangles) are coloured by sex, age (quartile bins) and sequencing depth (mapped reads; quartile bins). Only 5,667 unmatched controls in which all six events could be quantified are shown. **C)** False-positive rate (FPR) among unmatched whole-blood controls when aggregating combinations of one to five of the signature sites. For each combination, weighted Δlogit-PSI values are summed across those sites, and a false positive is defined as a score higher than the score of the lowest scoring n.64_65insT individual for that combination. Boxplots show medians and interquartile ranges (IQRs) across combinations of that number of sites; whiskers extend to the most extreme points within 1.5 × IQR. Points are jittered horizontally.

**Supplementary Figure 7:**
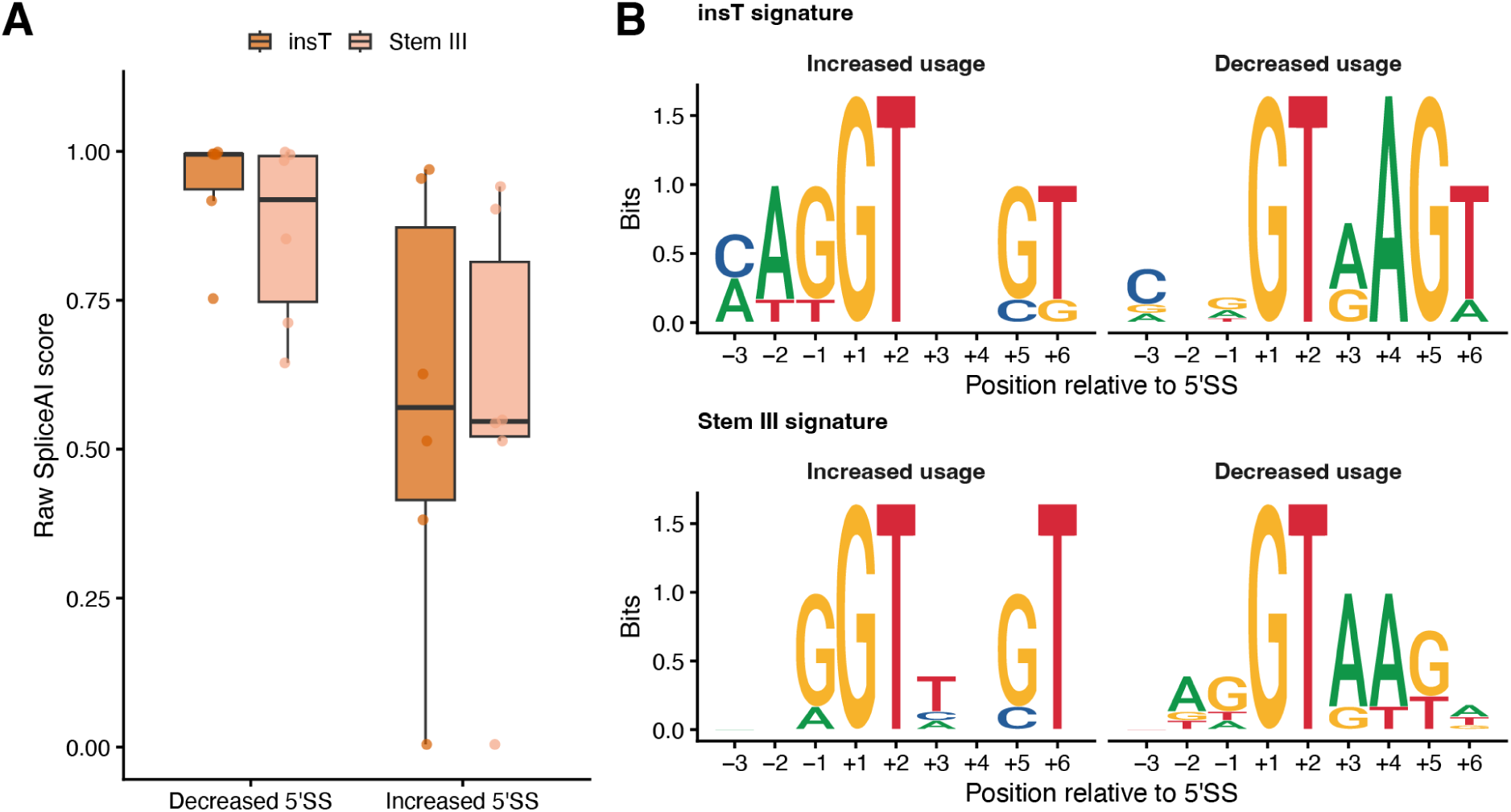
Characterisation of T-loop and Stem III signature splice event 5’SS motifs. **A)** Box plots showing raw spliceAI scores of the decreased 5′SS site and the increased 5′SS for the insT signature events (n=6) and stem III signature events (n=6). Boxes indicate the interquartile range with median and outliers shown. **B)** Position weight matrix sequence logos for the 5′ splice-site donor sequences associated with the insT and Stem III signature A5SS events. Events were stratified according to if ReNU cases show increased or decreased usage of the 5’SS.

**Supplementary Figure 8:**
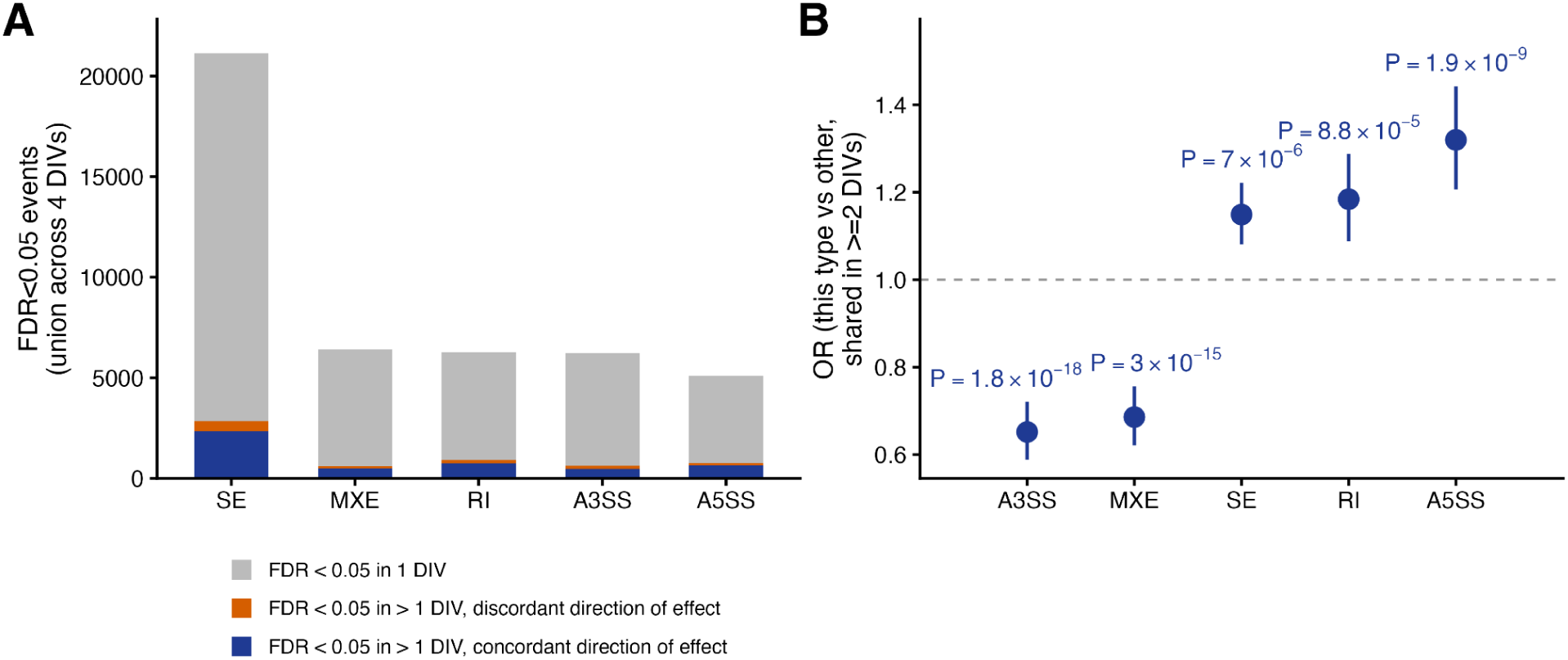
Reproducibility of aberrant splicing events usage across iPSC differentiation. **A)** Numbers of alternative splicing events significantly altered between ReNU and KOLF cells (FDR < 0.05) across the four stages of neuronal differentiation, stratified by rMATS event type. Events represent the union of significant events across DIV1, DIV5, DIV9 and DIV14 and are classified as significant at exactly one differentiation stage (grey), significant at two or more stages with discordant directions of effect (orange), or significant at two or more stages with a concordant direction of effect across all stages at which the event was significant (navy). **B)** Odds ratios (OR) with 95% confidence intervals showing the likelihood of each alternative splicing event type being shared across multiple stages of differentiation with concordant direction of effect, compared with all other event types (two-sided Fisher’s exact test).

## Notes

### Competing Interest Statement

N.W., S.J.S., and C.R. receive research funding from Biomarin Pharmaceutical

### Author Declarations

Secure access to the NGRL under project ID RR354 was provided by Genomics England, which delivers the NGRL in partnership with NHS England, and is wholly owned by the UK Department of Health and Social Care. The NGRL contains participants health data collected by the NHS as part of their care, along with samples and data from their participation in research, for which fully informed consent has been obtained. Genomics England has approval from the HRA Committee East of England Cambridge South (REC Ref 14/EE/1112).

### Summary of Updates

Correcting a spelling error in the author list

